# Establishing health-based Biological Exposure Limits for pesticides: a proof of principle study using Mancozeb

**DOI:** 10.1101/2020.05.12.20098939

**Authors:** Stefan Mandić-Rajčević, Federico Maria Rubino, Claudio Colosio

## Abstract

Pesticides represent an economical, labor-saving, and efficient tool for pest management, but their intrinsic toxic properties may endanger workers and the general population. Risk assessment is necessary, and biological monitoring represents a potentially valuable tool. Several international agencies propose biological exposure indices (BEI), especially for substances which are commonly absorbed through the skin. Biological monitoring for pesticide exposure and risk assessment seems a natural choice, but biological exposure limits (BEL) for pesticides are lacking.

This study aims at establishing equivalent biological exposure limits (EBEL) for pesticides using real-life field data and the Acceptable Operator Exposure Level (AOEL) of mancozeb as the reference.

This study included a group of 16 vineyard pesticide applicators from Northern Italy, a subgroup of a more extensive study of 28 applicators. Their exposure was estimated using “patch” and “hand-wash” methodologies, together with biological monitoring of free ethylene-bis-thiourea (ETU) excretion in 24-hour pre- and post-exposure urine samples. Modeling was done using univariate linear regression with ETU excretion as the dependent variable and the estimated absorbed dose as the independent variable.

The median skin deposition of mancozeb in our study population was 125 μg, leading to a median absorbed dose of 0.9 μg/kg. The median post-exposure ETU excretion was 3.7 μg. The modeled EBEL for mancozeb was 148 μg of free ETU or 697 μg of total ETU, accounting for around 75% of the maximum theoretical excretion based on a mass balance model. Although preliminary and based on a small population of low-exposed workers, our results demonstrate a procedure to develop strongly needed biological exposure limits for pesticides.

## 1. Introduction

Pesticides represent a fundamental component of modern agriculture, and without their use, a significant production loss would be inevitable, in particular in developing countries (Cooper and Dobson, 2007; Oerke and Dehne, 2004). However, their toxic properties, biological activity, and weak specificity for target organisms may endanger applicators, farmers, the general population, and the environment (Mostafalou and Abdollahi, 2013; Parrón et al., 2011). For this reason, the assessment of all the risks consequent to their use is a necessary step (Rubino et al., 2012). Risk assessment of pesticides encompasses two main phases; the first one, the *“pre-marketing phase,”* is when all the key information regarding the chemical, physical, and toxicological properties of active ingredients are collected to establish the health-based limits for the general population and workers. In the European Union, the Acceptable Operator Exposure Level (AOEL) is defined as the dose of an active substance, expressed as milligrams per kilogram of body weight, that a worker can absorb during a workday, without an unacceptable risk of long term health effects (EC, 1991; Mandic-Rajcevic et al., 2013; Regulation, 2009).

Health risk assessment of exposure to noxious chemicals is based on the relationship between the individual’s estimated exposure and the established exposure limit, which is derived from the dose-effect relationship (Lebailly et al., 2008; Vitali et al., 2009). For workers in agriculture, especially in open-field farming where the prevalent route of exposure is the skin, personal exposure monitoring is usually based on the interception or removal techniques that allow reconstructing exposure through the dermal route (Lebailly et al., 2008; Pierre et al., 2010).

Biological exposure monitoring is the measurement of pesticides or their biotransformation products found in the body, or eliminated therefrom, appropriate time after exposure (Maroni et al., 2000). This strategy is suitable for measuring workers’ absorbed doses independently of the route of exposure, absorption variation, and timing (Colosio et al., 2002; Fustinoni et al., 2014). Several national and international health protection agencies establish biological exposure limits for various chemicals for which the main route of exposure is inhalation, such as industrial solvents. The American Conference of Governmental Industrial Hygienists (ACGIH®) develops Biological Exposure Indices (BEI®) based on their relationship with the workplace airborne Threshold Limit Values (TLV®) or corresponding adverse health effects (ACGIH, 2004). In Germany, the MAK commission recommends the Maximum Workplace Concentrations (MAK values) in the air and Biological Tolerance Values (BAT values) in blood or urine (Forschungsgemeinschaft), 2017). The EU Scientific Committee on Occupational Exposure Limits (SCOEL) develops and publishes health-based Occupational Exposure Limits (OELs) and, when possible, the corresponding biological limit values (BLVs) (Bolt and Thier, 2006).

In public health settings, the use of biomonitoring equivalents has been proposed to estimate the amount of a chemical or metabolite in a biological specimen that is consistent with an existing exposure guidance value such as a tolerable daily intake or a reference dose (Hays et al., 2008; Hays and Aylward, 2009). In occupational settings, a Biological Exposure Limit is based on the direct proportionality between the airborne concentration of the compound and its concentration, or that of its metabolites, in suitable biological specimens (SCOEL, 2013). The ACGIH and the SCOEL define the respective BEI and BLV similarly as follows: *“…the levels of determinants which are most likely to be observed in specimens collected from healthy workers who have been exposed to chemicals to the same extent as the workers with inhalation exposure at the TLV (or OEL in case of SCOEL)”* ((SCOEL), 2014; ACGIH, 2004).

Unfortunately, health-based biological exposure limits for pesticides are still lacking, rendering the interpretation of biological monitoring results extremely difficult. The AOEL for pesticides is a health-based occupational exposure limit expressed as an internal dose. Therefore, starting from the AOEL, it should be possible to establish an Equivalent Biological Exposure Limit (EBEL). An EBEL can be defined by building on the ACGIH’s relationship of the BEI to the TLV and the EU-SCOEL’s relationship of the BLV to the OEL. The definition needs to take into account the intrinsic difference between the TLV/OEL and the AOEL. The TLV/OEL are airborne concentrations that can be transformed into a systemic dose by assuming an individual’s ventilation rate and exposure time. On the contrary, the AOEL is already a systemic dose. A definition of the EBEL that takes into account this important difference would be: **”…levels *of determinants of exposure to a specific pesticide which are most likely to be observed in biological specimens collected from healthy workers who have absorbed, through all routes of exposure, a total dose of the pesticide equivalent to its AOEL”***.

**This study aims to propose a procedure to establish a “proof of principle” EBEL using real-life field data and AOEL of mancozeb as the guidance value**.

## 2. Materials and Methods

Mancozeb is a fungicide that has been used in various crops for decades. Chemically it is a polymeric 1:1 mixture of the complexes of ethylene-bis-dithiocarbamate with Manganese and with Zinc. Ethylene-bis-thiourea (ETU) is its primary urine metabolite, which is produced from mancozeb in approximately. 1:2 molecule ratio. ***Figure 1*** shows their chemical structures, composition, and molecular weights, as reported from the PubChem database. ETU can be measured in urine with sensitive and selective microanalytical techniques. Elevated post-exposure urine levels of ETU highly correlated with the estimated occupational exposure have been found previously (Colosio et al., 2002; Mandic-Rajcevic et al., 2018). Mancozeb is rapidly and completely excreted: according to its authorization report, 50% is excreted within one day and >95% within four days (SANCO, 2009).

**Figure 1.**
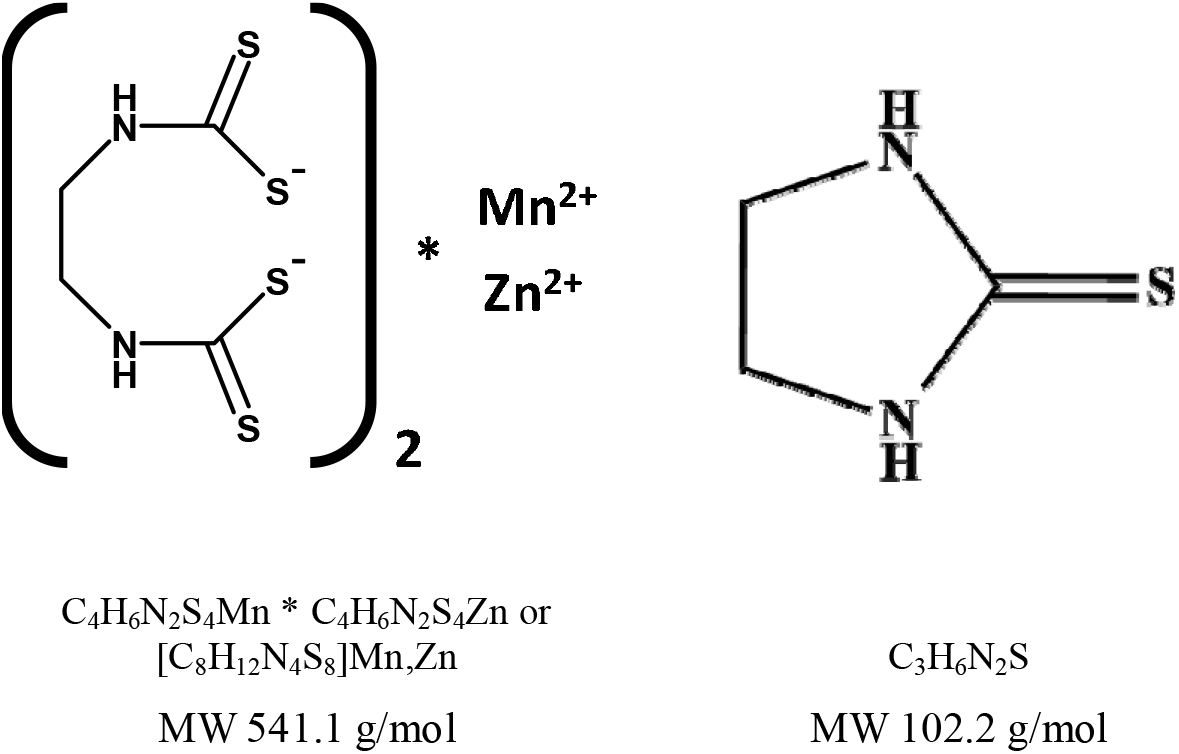
Mancozeb (left) and its metabolite ethylene-bis-thiourea (right)

In 2011, a group of pesticide applicators spraying mancozeb on vineyards in Lombardy (Northern Italy) were monitored during their workdays. They performed regular activities of mixing and loading, application, maintenance and cleaning of their equipment. Workers were identified and contacted through agricultural support organizations and local health service of the provinces of Mantova and Pavia. Employers and their workers were invited to a presentation of the study protocol, where the planned data and sample collection was explained to them, and they chose whether they wish to participate in the study. The workers officially applied for participation by signing the study informed consent form. The Ethics Committee of the San Paolo Hospital (University of Milan teaching hospital) approved the study in February 2011.

Three levels of data collection were included in our study protocol:

a. a data collection checklist developed specifically for pesticide exposure studies which includes a large number of variables which have been identified to influence exposure (Mandic-Rajcevic et al., 2015);
b. skin exposure assessment using the “patch” and “hand-wash” techniques according to a modified protocol of the Organization for Economic Co-operation and Development guidelines (Mandic-Rajcevic et al., 2018; OECD, 1997);
c. biological monitoring by the collection of 24-hour pre- and post-exposure urine samples. The post-exposure urine sample collection started at the end of the application and lasted for 24h.

To improve the assessment of mancozeb deposition on the skin and the estimate of the absorbed dose, we examined the analytical results to identify and treat outliers. Outliers that could influence the estimate of the skin dose were identified and treated using the modified Z score according to our previously proposed and published procedure (Mandić-Rajčević and Colosio, 2019; Mandic-Rajcevic et al., 2019). This processing improved the correlation between the estimated skin exposure and absorbed dose and the ETU levels found in post-exposure urine samples.

### 2.1. Study sample

Twenty-eight workers applying mancozeb on 38 workdays participated in the original study. Workers who reported any occupational exposure to ethylene-bis-dithiocarbamates in the 15 days before the study day, had any missing of “patch” or “hand-wash” sample, or missing or unreliable 24-hour urine sample (verified by urine and creatinine quantity) were excluded from this study. Having in mind that the application often takes several days in specific periods to fully treat the farmers’ estate, and the treatment is often followed by re-entry work, which can result in additional exposure, the safest option was to have a longer non-treatment window before biological monitoring. After applying the exclusion criteria, 16 workers and their workdays were included in the study.

The in-field study team (Authors: SMR and FMR) gathered the information and filled in the data collection sheet from the workers (sociodemographic characteristics), from the supervisors (farm characteristics), and by observing and documenting (photos) the actual in-field conditions (various exposure determinants, personal protective devices use).

### 2.2. Laboratory analyses

During the application in the field, the workers had limited to no chance of urinating. During the 24-hour post-exposure urine collection, the container was kept in a dark and cool place or a domestic refrigerator. ETU in different kinds of samples (pad, handwash, and urine) was measured by liquid chromatography-mass spectrometry in an Acquity UPLC system (Waters, Milford, MA, USA) coupled with a triple quadrupole Waters TQD mass spectrometer. A detailed description of the laboratory analyses can be found in our previously published article (Mandic-Rajcevic et al., 2019). ETU determinations in urine samples did not include prior urine hydrolysis and thus measured only the free ETU (Fustinoni et al., 2005; Jones et al., 2010; Sottani et al., 2003).

In short, urine samples (2ml) were diluted with water (1 ml), spiked with ^2^H_4_-ETU, and purified using diatomaceous earth column (ChemElut® 3ml unbuffered, Varian, Poole, UK) (Jones et al., 2010). The calibration curve (constructed with a pool of urine of no-smoking subjects) was linear in the range 2.5-100 μg/l. The limit of detection (0.1μg/l) was defined as the amount injected (from an extracted urine sample) that gave a signal equivalent to three times the baseline noise. Clothes and skin pads samples (8×12.5cm) were spiked with ^2^H_4_-ETU, inserted in a polypropylene tube, and desorbed with 8 ml of water, vortexed for 10 minutes, centrifuged and an aliquot was injected onto UPLC after a suitable dilution factor with 0.1% formic acid solution. The calibration curve was linear in the range 1-50μg for clothes pads and 5-500ng for skin pads. Hand wash samples were obtained with 25% isopropyl alcohol (500mL*2); an aliquot was centrifuged, diluted 1:20 in 0.1% formic acid (1ml), spiked with ^2^H_4_-ETU and finally injected in UPLC (3 μl). The calibration curve for these samples was linear in the range of 0.2-4 mg. For all the type of samples two quality control were run after every ten samples; the recovery was at least > 77% for urine samples and > 86 % for the other type of samples; percent CV was always lower than 15%.

### 2.3. Absorbed dose assessment

The absorbed dose was calculated from the measured skin exposure, taking into account the surface of the pads and extrapolating on the surface of the skin represented by the pad, using the dermal absorption coefficient of 0.24% established for mancozeb during the authorization process (SANCO, 2009). The dermal absorption coefficient had been established for a standard 8-hour exposure, while the duration of the workday of our study participants varied from several hours to up to 10 hours, and they washed their hands from 1-7 times during the workday. Thus, we deemed necessary, including in the calculation of the absorbed dose the real duration of exposure. This was accomplished by using the first-order kinetic equation according to our previously proposed and published method (Mandic-Rajcevic et al., 2019). Calculations were performed on analytical results converted from mass units (micrograms or nanograms) to molecular units (nanomoles). Final results are expressed in micrograms or nanograms, for homogeneity with the mass units employed for the AOEL, and to retain the correct number of digits for easy reading. The AOEL saturation represents the ratio between the actual dose for each worker (in mg/kg of bw) and the AOEL (in mg/kg bw). Considering that AOEL is a limit value, we express this ration as the “saturation” of the limit and in %. In case the ratio is over 1 or the saturation is over 100%, the worker is considered overexposed.

### 2.4. Statistical analyses and establishment of an EBEL

The measurements of ETU acquired in the course of field studies were not normally distributed; therefore, median values, with minimum and maximum values between parentheses, are reported in text and tables. Categorical variables are presented as the number of occurrences and percent in text and tables.

EBEL was established using univariate linear regression. Since exposure data were not normally distributed, the absorbed dose and 24-hour post-exposure urine ETU levels were log-transformed. The distribution of the log-transformed data was verified by visualizing the data and using the Shapiro-Wilk test. The relationship between the absorbed dose (of mancozeb; independent or explanatory variable) and the excreted metabolite (ETU measured in urine; dependent or outcome variable) was modeled on log-transformed data.

The Equivalent Biological Exposure Limit for mancozeb was predicted as the amount of ETU corresponding to that most likely to be found for the absorbed dose of 0.035 mg/kg of body weight (the AOEL for mancozeb) using the model equations calculated in the previous step (SANCO, 2009).

A recent article demonstrated that free ETU in the urine might represent only approximately 20% of the total ETU measured after hydrolysis (Ekman et al., 2013). Thus, to use only free ETU as the dependent variable may represent a potential weakness in this approach. To improve the estimated ETU excretion, we have calculated the value of “Total ETU” by employing the equation reported in the paper by Ekman et al. (2013, ***Equation 1***).

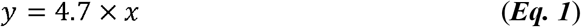

Our models thus report both “Free ETU” and “Total ETU” as dependent variables.

### 2.5. Excretion simulation and mass balance of the predicted EBEL

The reliability of prediction of ETU excretion according to the proposed model was assessed by comparing the EBEL simulated for “Free ETU” and for “Total ETU” in a 70-kg worker who has absorbed a dose of mancozeb corresponding to the AOEL (dose saturation) with the theoretical excretion of ETU calculated with a stoichiometric mass balance.

## 3. Results

### 3.1. General characteristics

Sixteen male workers were included in this study, of whom 5 (31%) used open, and 11 (69%) closed and filtered tractors. ***Table 1*** summarizes workers’ characteristics and the characteristics of the workday and personal protective equipment of the study population. There was no statistically significant difference in age, height, weight, and experience between the workers using the two kinds of tractors. However, workers using open tractors applied mancozeb in smaller areas, and this difference was statistically significant (p = 0.013).

**Table 1.** Characteristics of the study population and workdays included in the development of the model.

|  | [ALL]<br>N=16 | Open<br>N=5 | Filtered<br>N=11 |
| --- | --- | --- | --- |
| Sex: Male | 16 (100%) | 5 (100%) | 11 (100%) |
| Age | 44 (32-60) | 43 (36-47) | 44 (32-60) |
| Height | 174 (162-190) | 178 (173-184) | 172 (162-190) |
| Weight | 83 (62-120) | 100 (75-120) | 80 (62-100) |
| Experience (years) | 15 (4-35) | 20 (4-24) | 15 (6-35) |
| Area (ha) | 7 (1-20) | 2 (1-10) | 7 (3-20) |
| Coverall: |  |  |  |
| None | 3 (19%) | 0 (0%) | 3 (27%) |
| Multy-use | 2 (12%) | 2 (40%) | 0 (0%) |
| Mono-use | 11 (69%) | 3 (60%) | 8 (73%) |
| Gloves Available: |  |  |  |
| No | 1 (6%) | 0 (0%) | 1 (9%) |
| Yes | 15 (94%) | 5 (100%) | 10 (91%) |
| Respiratory mask: |  |  |  |
| No | 4 (25%) | 1 (20%) | 3 (27%) |
| Paper | 2 (12%) | 1 (20%) | 1 (9%) |
| Filter | 10 (62%) | 3 (60%) | 7 (64%) |

### 3.2. Exposure and absorbed dose

***Table 2*** summarizes mancozeb deposition on the skin, absorbed doses, AOEL saturation, urine volume, creatinine, and mancozeb levels before and after the exposure. The median deposition of mancozeb on the skin was 2.4 μg (0.1 - 529.9 μg), while the median deposition on the hands was 123.9 μg (19.6 - 4724.2 μg), resulting in a median total deposition of 125.8 μg (19.7 - 4747.2 μg). Higher skin deposition of mancozeb was measured in open tractor workers compared to closed and filtered tractor workers, 240.8 μg compared to 69.1 μg, respectively.

**Table 2.**
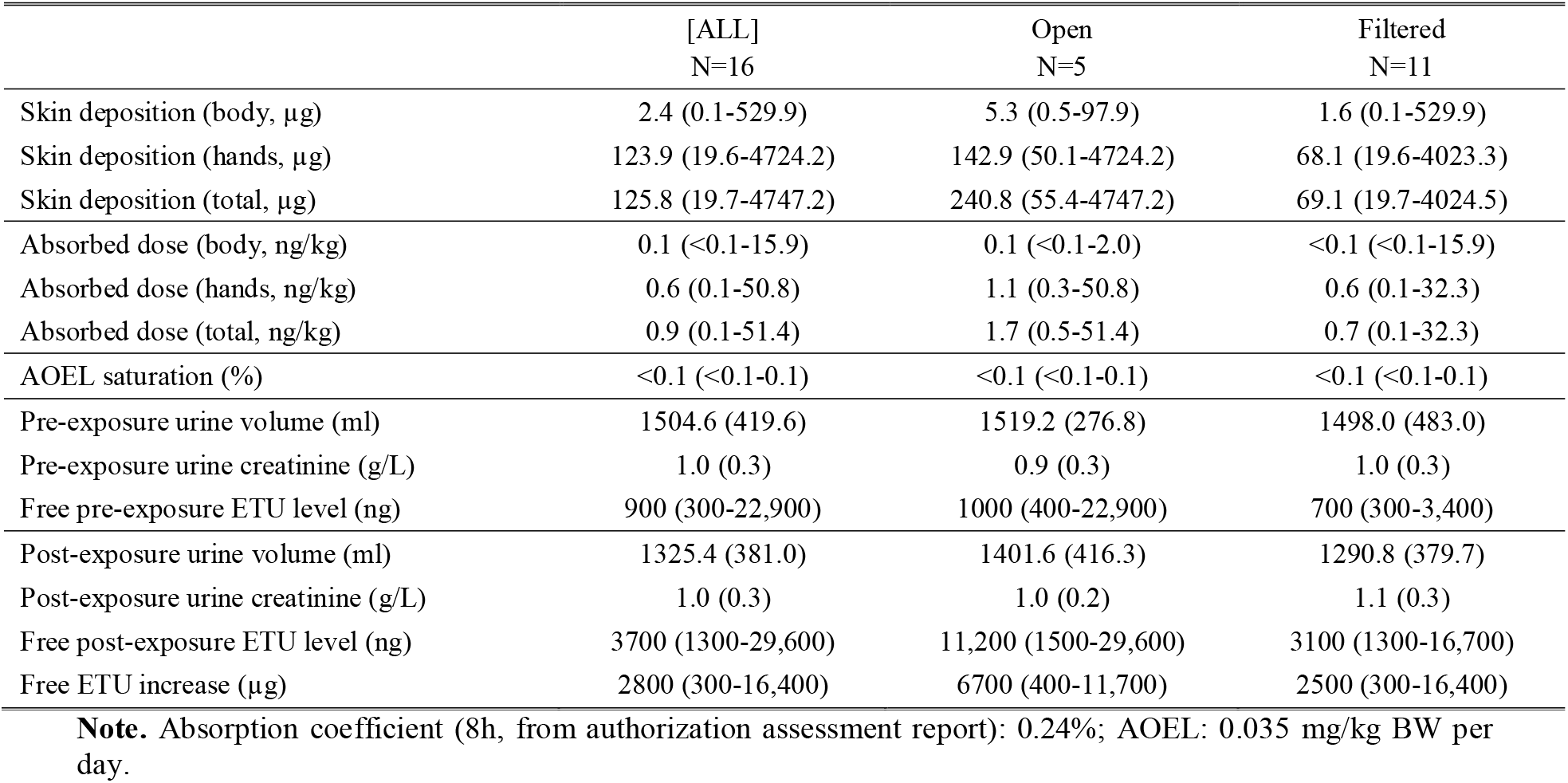
Absorbed dose, AOEL saturation, pre- and post-exposure urine volumes, creatinine, and free ETU levels.

Considering the 8-hour absorption coefficient of mancozeb of 0.24%, and workers’ weights, the estimated absorbed dose from body exposure was 0.1 ng/kg (<0.1 – 2.0 ng/kg), while the median absorbed dose from hands exposure was 0.6 ng/kg (0.1 – 50.8 ng/kg), resulting in a total absorbed dose of 0.9 ng/kg (0.1 – 51.4 ng/kg). Equivalent to the skin deposition, the median total absorbed dose in open tractor workers was higher than that of closed and filtered tractor workers, 1.7 ng/kg compared to 0.7 ng/kg.

Taking into account the AOEL for mancozeb of 0.035 mg/kg, the saturation of AOEL of our workers was less than 1%, or differently, even our highest exposed worker was 1000 times less exposed than the daily limit for mancozeb.

### 3.3. Biological monitoring

The median pre-exposure urine volume, creatinine, and free ETU levels were 1504.6 ml, 1.0 g/L, and 900 ng, respectively. There was no statistically significant difference between the open and closed and filtered tractor workers. The median post-exposure urine volume, creatinine, and free ETU levels were 1325.4 ml, 1.0 g/L, and 3700 ng, respectively. The median values of free ETU were 11,200 and 3100 ng for the open and closed and filtered tractor workers, respectively. Although notable, this difference was not statistically significant (p = 0.183). Finally, the median pre- to post-exposure difference in urine free ETU levels was 2800 ng, with a higher difference of 6700 ng in open, and lower difference of 2500 ng in closed and filtered tractor workers.

***Figure 2*** shows the free ETU levels of the individual workers before the exposure (left) and after the exposure (right). Symbols mark the type of tractor used, lines connect each workers’ pre- and post-exposure urine ETU levels, and the color of the lines represents the absorbed dose. Although the levels of exposure were low relative to the AOEL, data confirm more significant increases in the ETU levels in workers with higher absorbed doses.

**Figure 2.**
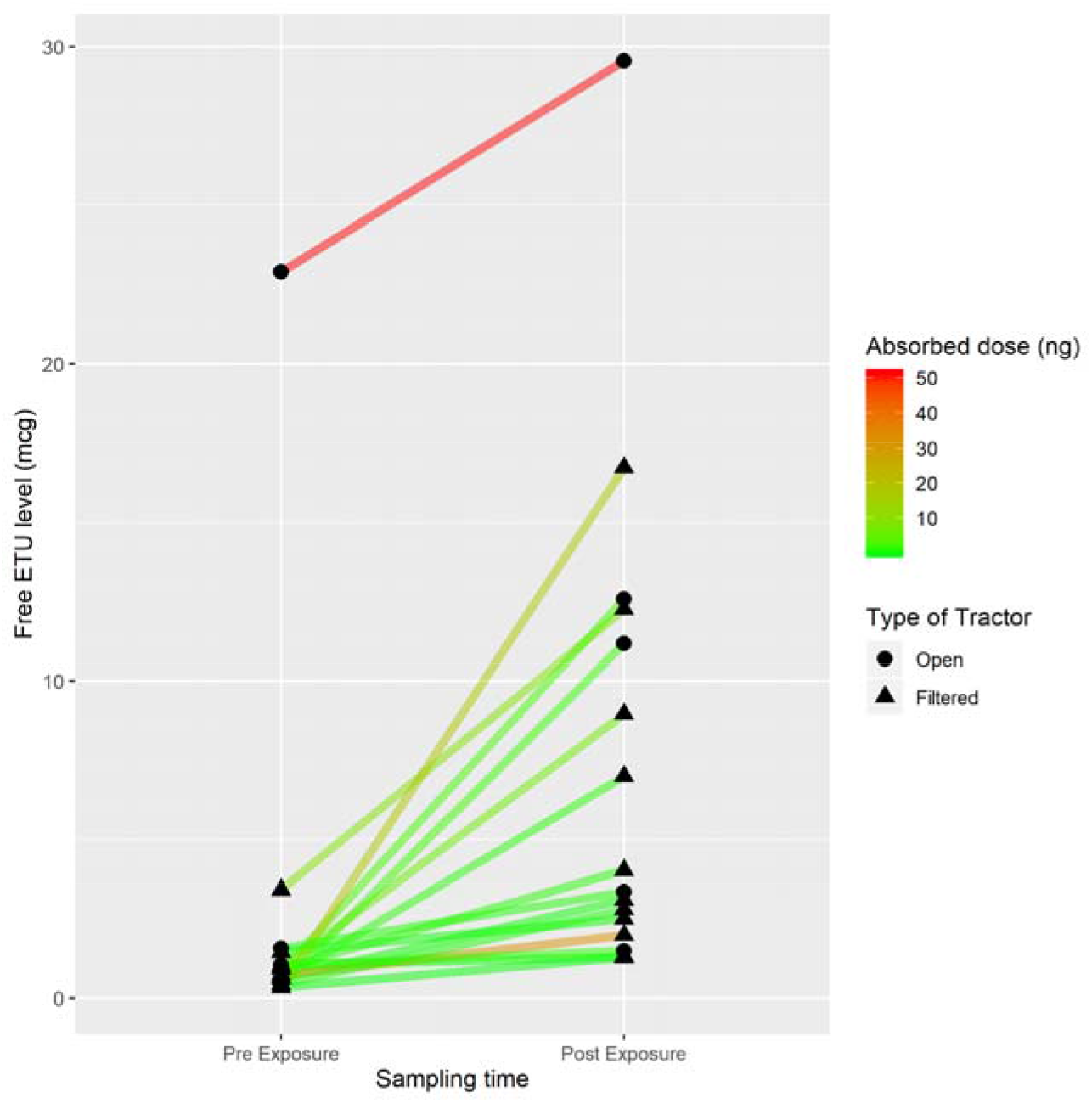
Individual increase in the urine ETU level depending on the absorbed dose.

### 3.4. Establishment of an EBEL

***Figure 3*** shows the two linear regression curves that estimate the urine levels of “free ETU” and of “Total ETU,” depending on the absorbed dose of mancozeb in our workers. “Total ETU” is calculated according to the correction for conjugated ETU suggested by Ekman et al. (2013) (Ekman et al., 2013).

**Figure 3.**
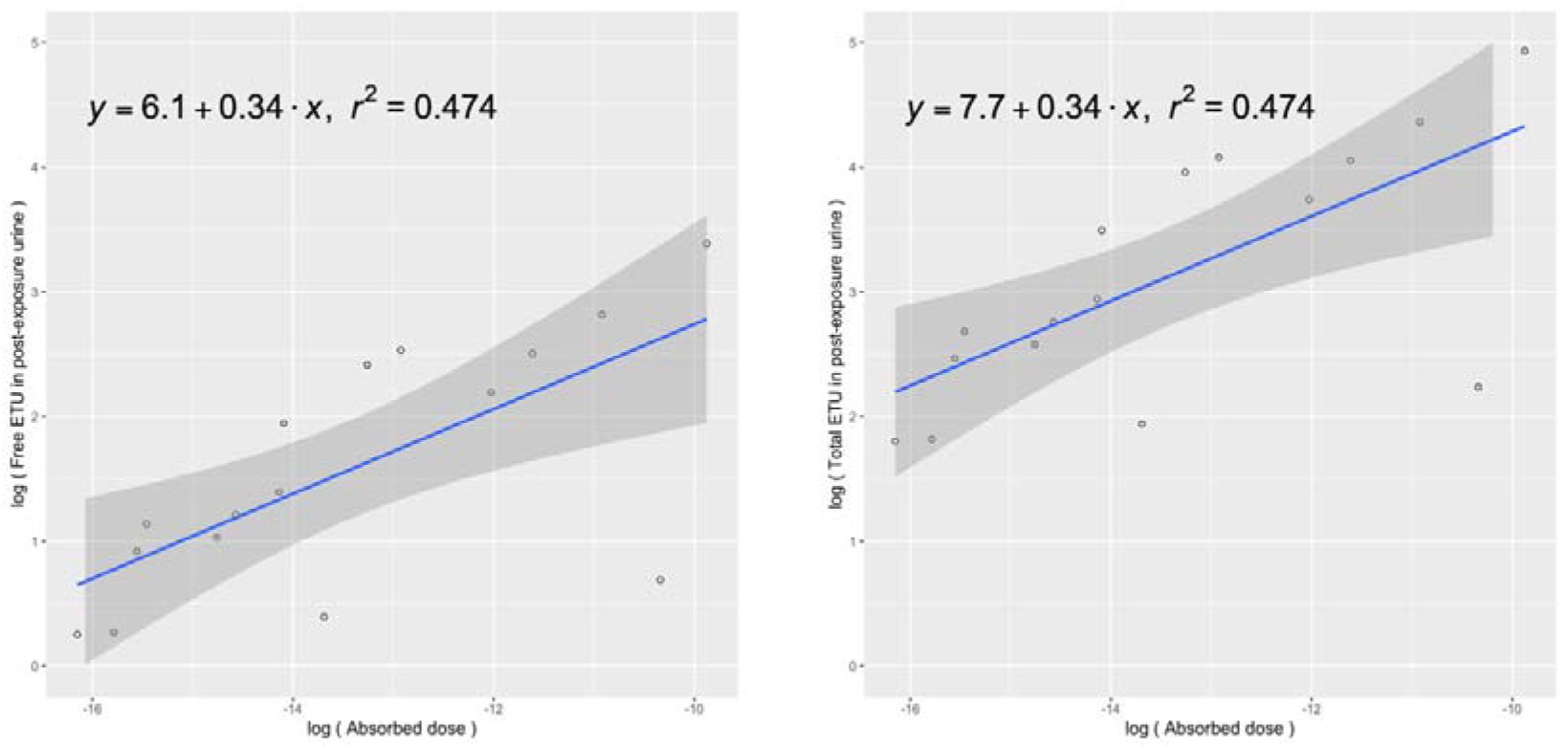
Linear regression curve estimating the free ETU (left) and total ETU levels (right) in urine depending on the absorbed dose (all values are log-transformed)

The linear regression equations are, respectively:

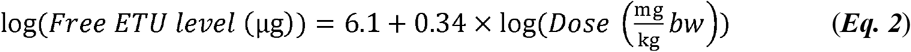

And

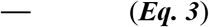

Using the “Free ETU” model for prediction (***Equation 2***), for a worker who absorbed, through all routes of exposure, the dose equivalent to the AOEL of 0.035 mg/kg bw/day, the predicted 24-hour post-exposure urine free ETU level would be 148 μg (95% CI: 18 - 1,221 μg^)^.

Using the “Total ETU” model for prediction (***Equation 3***), for a worker who absorbed, through all routes of exposure, the dose equivalent to the AOEL of 0.035 mg/kg bw/day, the predicted 24-hour post-exposure equivalent ETU level would be 697 μg (95% CI: 84 - 5,742 μg^)^.

According to the reported regression equations, the resulting EBEL for mancozeb is 148 μg of “free ETU” in 24-hour post-exposure urine, or 697 μg of “Total ETU” (after hydrolysis) in 24-hour post-exposure urine.

### 3.5. Excretion simulation and mass balance of the predicted EBEL

Excretion simulation of biomonitoring results according to a mass balance model has been performed for a theoretical (standardized) 70-kg subject. The calculation, according to the AOEL of 0.035 mg/kg bw results in a maximum allowable body burden of approximately 2.45 mg of mancozeb. Taking into account the molecular mass of mancozeb, of ETU and considering that one mole of mancozeb affords two moles of ETU, the corresponding total amount of ETU 0.92 mg. When the predicted amount of free ETU based on our model (***Equation 1***) of 0.15 mg is compared to the theoretical value of 0.92 mg, it represents approximately 15%. However, recent literature suggests that a further amount of ETU can be measured in urine when the pre-analytical sample preparation step includes an acid treatment that hydrolyzes yet undetermined bound forms of ETU (Ekman et al., 2013) (see ***Material and Methods***). When the correction factor they proposed is applied to our data, the predicted amount of total ETU according to Equation 2 is 0.7 mg or approximately 75% of the theoretical ETU excretion, corresponding to the expected 24-hour excretion. The predicted amount corresponds to 10 μg/kg bw ETU excretion in 24-hour post-exposure urine.

## 4. Discussion

The need for food produced by the agricultural sector is ever growing with the growth of the population. The lower yield achieved by some regions, such as Sub-Saharan Africa and the loss of almost 25% of food calories between the field and fork (Flood, 2010) indicate that the use of pesticides around the world will not be declining anytime soon. Controlling pesticide applicators’ working conditions by environmental monitoring is hindered by the complexity of available methods, their price, but also poor reproducibility due to extreme variations in the climatic conditions, work modalities, available equipment, and use/nonuse/misuse of personal protective devices characterized by different levels of quality (Arbuckle et al., 2002; Mandic-Rajcevic et al., 2018, 2015). Biological monitoring, although more convenient, relies on the availability of biological exposure limits, which exist for several substances used in industrial settings but are lacking for pesticides. Considering that almost one-third of the world’s population works in the agricultural sector, commonly in poor working conditions, the need to propose biological exposure limits for pesticides is underlined. This study aimed at finding a strategy to derive equivalent biological exposure limits for the monitoring of workers who use pesticides in agricultural, public health, and other occupational settings.

Our study included only 16 workers, 5 working with open and 11 working with closed and filtered tractors. The main noticed difference between the two groups was the smaller area covered by open tractors, which is inherent to the application modality. The size of our study population is similar to other real-life studies of pesticide applicators. Most studies conducted in Europe include between 10 and 15 workers, although even smaller numbers have been reported (Aprea, 2012; Rubino et al., 2012; Tsakirakis et al., 2014). Our study results are based on a group of low-exposed and well-protected workers, which is common for the North of Italy, and even Europe, but quite different from exposure in some developing countries.

All workers supplied data for the data collection sheet, which was designed to collect all the information about the workers, workday, determinants of exposure, which have been cross-checked and filled by a member of our field team. Detailed information about work characteristics and their combined influence on exposure levels has been published previously and is not included in this manuscript to avoid repetition (Mandic-Rajcevic et al., 2015). The median skin deposition of mancozeb in our workers was 125 μg, while the median absorbed dose was 0.9 ng/kg of body weight. The low exposure (skin deposition) to pesticides seen in our group confirms similar findings in European studies (Mandic-Rajcevic et al., 2015; Tsakirakis et al., 2014). The modality of application, namely the use of closed and filtered tractors by the majority of our workers, also reduces the exposure to the active substance (Arbuckle et al., 2002; Hines et al., 2011). The use of adequate spraying equipment, as well as personal protective devices, is commonly underlined as the primary way of exposure prevention in agricultural pesticide applicators (Damalas and Koutroubas, 2016). In our study, only skin was considered, as it is the main route of exposure to pesticides accounting for 99% of exposure in open field farming (Lebailly et al., 2008). In a more experimental setting, both inhalatory and dermal monitoring could be applied to quantify the exposure, having in mind the increased burden on the study participants and the reduction in the number of subjects monitored. Each approach has its advantages and disadvantages and is connected with a tradeoff.

The absorbed dose was more than 1000 lower than the acceptable operator exposure level, indicating there is no health risk for our workers. For mancozeb, considering the dermal absorption coefficient of mancozeb of 0.24% (SANCO, 2009), the low absorbed dose was also to be expected.

Exposure to ethylene bisdithiocarbamates (EDBCs) of which mancozeb is a member can be considered ubiquitous, most likely due to residues in food and wine, and measurable levels have been found even in the general population (Corsini et al., 2005). The median level of 0.9 μg found in pre-exposure urine samples of our population falls in the range of the general population. Nevertheless, one of our workers had the pre-exposure free ETU level in urine of 22.9 μg, which would indicate occupational exposure, although he declared not to have used EDBCs in the previous 15 days. Post-exposure urine samples had a median level of 3.7 μg of free ETU, with a much higher increase in open tractors (6.7 μg increase) compared to closed and filtered tractors (2.5 μg). In the case of post-exposure urine, the worker with the highest level was the one with the highest pre-exposure urine free ETU level, indicating that it was not the work activity of the monitored day, which caused the exposure. Nevertheless, ETU was once again a reliable indicator of occupational exposure to EDBCs, in our case, to mancozeb. In our study, we have excluded workers with no increase of ETU after work, as this would indicate higher exposure from uncharacterized sources. Taking into account other sources of exposure would also be important but were unable to reliably quantify and insert these levels of exposure into our model. Having in mind that our attempt is the first step towards equivalent biological exposure limits for pesticides, additional uncertainty would decrease its value. This study also did not include a food consumption questionnaire, however, a cognate study had been carried out with a replicate diet and questionnaire to assess non-occupational exposure vs. occupational exposure to conazole fungicides, and the diet dose was less than 1% of the total dose of a worker working even in the conditions of best personal protection (Kennedy et al., 2015).

Our approach proposed an EBEL for mancozeb of 0.15 mg of free ETU or 0.7 mg of total ETU in urine, depending on the method used for laboratory measurement of ETU in urine samples. These values represent around 75% of the maximum theoretical amount, which could be excreted by a 70 kg worker who has absorbed, through all routes of exposure, the amount of mancozeb equivalent to the AOEL. Having in mind the literature which suggests between 50-75% of ETU would be excreted during the first 24 hours, we compare our modeled results to this figure. This comparison demonstrates the feasibility of our approach to establishing health-based equivalent biological exposure limits for pesticides, similar to the use of biomonitoring equivalents, but in an occupational setting (Hays et al., 2007). Considering the proposed EBEL, even our most exposed worker would be at the level of 20% of the EBEL, while most of the other workers would be at or below 3% of the EBEL. Therefore, EBEL could prove a useful screening tool for pesticide applicators, the first-tier assessment through only one sample, and one laboratory analysis whether the workers are close to being at risk for their health.

It should be considered that mancozeb might represent a perfect prototype of an active substance for which an EBEL can be established, as it is not accumulated, it is quickly metabolized and excreted mostly in urine with a half-life of 1 day (SANCO, 2009). This indicates that the 24-hour window of post-exposure urine collection applied in our study could be appropriate for this active substance. Although we have taken into account both the “free” ETU and “total” ETU, the study, which has discovered the existence of these two compartments for the excretion of ETU found the same toxicokinetic characteristics in both forms (Ekman et al., 2013). We, therefore, do not expect relevant differences in excretion times. Additional attention would be necessary for active substances that accumulate in the body or are excreted during longer periods, so the feasibility of developing an EBEL should be assessed on a case to case basis. Since the EBEL is developed for a specific active substance, using field studies and authorization data on exposure and excretion, its use in the current form is limited to the specific active substance. Nevertheless, an EBEL could be developed for a group of substances, such as EDBCs, with a common metabolite. A common ETU value could represent a common EBEL, or the EBEL could be represented in “mancozeb equivalents.” Another extension of an EBEL would be for risk assessment of exposure to mixtures of pesticides of known proportions. The saturation of an EBEL in the mixture could indicate, through proportion, the levels of exposure to other substances in the mixture. A similar approach has already been tested for Tebuconazole with promising results (Mandic-Rajcevic et al., 2015).

Our approach was developed in a group of male workers in a favorable situation of low occupational risk. The low exposure/risk introduces an unavoidable degree of uncertainty in the calculation, due to extrapolating the trend of exposure and excretion over two orders of magnitude. The assessment of the internal dose from available body deposition data relied on dealing with several sources of variability that include different deposition in the body areas, different duration of pesticide permanence on the skin, and a generic number for skin absorption coefficient (Mandić-Rajčević and Colosio, 2019; Mandic-Rajcevic et al., 2019; SANCO, 2009). Technical improvements in each of these aspects will increase the accuracy of the approach and the assessment. Some decisions to reduce the uncertainty in the quantification of the proposed EBEL, such as the exclusion of workers for whom it was impossible to quantify other relevant sources of exposure, or those who did not collect the expected volume of urine, have increased the uncertainty in the extrapolation and generalization to mancozeb applicators in general.

Correction by body weight is a useful tool to allow more accurate risk assessment, especially in children where the body weight can vary by a factor of four and even more, depending on the age (Valcke and Bouchard, 2009). The proposed EBEL translates to 10 μg per kg of body weight of ETU in the 24-hour post-exposure urine, which can be applied in cases of significant body weight variation, not often seen in male workers, but is possible between males and females in agriculture. Although all participants in our study were male, the female workforce is growing and females represent over 40% of agricultural workers in developing countries, and they may be involved in the use of pesticides and protection of crops (“Women in agriculture | Reduce Rural Poverty | Food and Agriculture Organization of the United Nations,” n.d.). The inclusion of critical exposure scenarios, such as the greenhouses, of an increased variability of population: the female in their fertile age, different ethnicities with inherent ethno-genomic and physio-anatomical characteristics, make necessary gaining more detailed knowledge on the ADME of pesticides and on their variability. Switching from the “average male worker” to more detailed metrics, such as relating the EBEL to the body mass, similar to AOEL, may improve its application where large inter-individual differences are expected in the working population.

Future studies should focus on improving both the exposure estimates derived from personal exposure monitoring, as well as the biological monitoring techniques. Advances in microanalytical techniques will benefit the latter due to the possibility of increasing molecular specificity in the measurement of metabolites (Ekman et al., 2013). Finding agricultural workers whose exposure and the consequent absorbed dose is closer to the AOEL would reduce the variability introduced by extrapolating over two orders of magnitude. When taking into account that exposure at the AOEL is considered safe even if a worker would be exposed every day for the duration of his life, volunteer studies could be planned to help derive valid biological exposure levels. As the design of the study moves from observational to experimental, additional attention is necessary to keep research within the boundaries of ethical behavior (London et al., 2010).

### Conclusions

Considering the importance of pesticide use of global agriculture, the number of workers exposed worldwide, and the dermal route as the main route of exposure, biological monitoring becomes the natural approach for risk assessment. The lack of biological exposure limits can be overcome by the development of equivalent biological exposure limits, based on real-life field exposure studies using environmental and biological monitoring in parallel and data from the authorization documents. In perspective, the establishment of equivalent biological exposure limits for active substances could become a part of the authorization process, ensuring affordable, simple, and reliable risk assessment of pesticide applicators later on.

## Data Availability

Data is available upon reasonable request.

## Acknowledgments

We acknowledge the support of the Italian Institute for Insurance of Occupational Diseases and Accidents (INAIL) – Session of the Region of Lombardy, which funded this study.5.

